# Adjuvant Recombinant SARS-CoV-2 Spike Protein Vaccine Immunogenicity After Prior mRNA Vaccine Doses

**DOI:** 10.1101/2024.11.07.24316865

**Authors:** Jeffrey M. Adelglass, Paul Bradley, Miranda R. Cai, Gordon Chau, Raj Kalkeri, Shane Cloney-Clark, Mingzhu Zhu, Zhaohui Cai, Mark Eickhoff, Joyce S. Plested, Raburn M. Mallory, Lisa M. Dunkle

## Abstract

**Background:** To support heterologous vaccine regimens, periodic SARS-CoV-2 revaccination required immunogenicity and safety data for adjuvanted protein-based vaccines following prior mRNA doses.

**Methods:** This phase 3, open-label study (2019nCoV-312/NCT05875701) enrolled participants who received one dose of ancestral SARS-CoV-2 protein-based vaccine (NVX-CoV2373) in an earlier study (2019nCoV-307/NCT05463068) after a primary series and possibly one additional dose of mRNA vaccine. In the current study, participants received an additional dose of protein-based vaccine (ancestral [n=104] or Omicron BA.5 [n=40]) at least 180 days after their previous study dose. The primary objective was demonstration of noninferiority of neutralizing antibody (nAb) titers induced by the dose in this study versus the first dose of NVX-CoV-2373 in the earlier study. Safety was also evaluated.

**Results:** A total of 144 participants were enrolled. The ratio of anti-Wuhan nAbs (GMT, IU/mL [95% CI]) at Day 28 after this study dose (ancestral 393.2 [318.0–468.2]) versus previous study dose (396.6 [328.7–478.6]) was 1.0 (0.8, 1.2), meeting noninferiority. The seroresponse rate difference between doses was 7.4% (95% CI = −1.2%–16.5%), also meeting noninferiority. Omicron BA.5 nAb titers suggest cross-protection against emerging variants. The nAb ratio at Day 28 between Omicron dose in this study (835.0 [597.1–1167.6]) versus previous study ancestral dose (436.0 [305.6–622.2]) was 1.9 (1.5–2.5), exceeding superiority criterion. Local and systemic reactions were similar between doses and strains in both studies.

**Conclusions:** A heterologous regimen of two adjuvanted recombinant spike protein-based vaccine doses after mRNA vaccination produced robust immune responses, exhibiting cross-reactivity to newer variants.

## INTRODUCTION

Since 2019, SARS-CoV-2 has evolved from a newly identified human viral pathogen responsible for a high degree of morbidity and mortality worldwide to a vaccine-preventable, endemic viral pathogen that remains a significant cause of excess morbidity and mortality.^1, 2^ During this evolution, vaccines based on multiple manufacturing platforms, both conventional and novel, were developed and deployed globally.^3–8^ With the acknowledgement that the virus has become endemic and mutates frequently, repeated vaccination appears to be necessary.^9–12^ Regulatory and public health policy responses have adopted the strategy of recommending periodic vaccination similar to that employed for influenza (the United States [US] Food and Drug Administration [FDA] and the World Health Organization [WHO]).

Following initial vaccinations with a two-dose primary series that demonstrated safety, immunogenicity, and protective efficacy, most vaccines were evaluated for immunogenicity as subsequent doses.^13, 14^ Development of the first-in-class adjuvanted, recombinant SARS-CoV-2 spike (rS) protein vaccine, NVX-CoV2373, followed the same path; however, due to limited data regarding its use as an additional dose, NVX-CoV2373 was initially limited by some regulators to use as a single additional dose, despite public health recommendations for multiple periodic doses.^15^ Vaccine recommendations also often indicated that vaccines based on different platforms should not be assumed to be interchangeable, thus limiting additional doses to homologous (matched vaccine) administration and not supporting heterologous (different vaccine) administration.^16^ The objective of this study was to demonstrate satisfactory safety and noninferior immunogenicity following first and second NVX-CoV2373 doses as part of a heterologous series including multiple doses of the mRNA vaccines that predominated in the US.

## METHODS

### Study design, population, and procedures

Adults were recruited into this Phase 3, open-label study from participants 18–49 years of age who had been enrolled in an earlier study (2019nCoV-307/NCT05463068),^17^ applying the same inclusion and exclusion criteria. In Study 2019nCoV-307, a single dose of NVX-CoV2373 – an adjuvanted, protein subunit vaccine based on the ancestral Wuhan strain of SARS-CoV-2 – was administered at least 6 months after the participants’ most recent previous mRNA vaccination (mRNA-1273, Moderna, or BNT162b2, Pfizer-BioNtech) or other SARS-CoV-2 vaccines.^17^ Dosing in Study 2019nCoV-312 similarly had to be administered at least 180 days after the dose in Study 2019nCoV-307 among participants who were dosed with mRNA vaccination.

In the current study (2019nCoV-312/NCT05875701), the first group enrolled received a dose of the ancestral strain–based vaccine (NVX-CoV2373), while a second group of participants, enrolled approximately 2 months later, received a single open-label additional dose of the Omicron BA.5 variant vaccine (NVX-CoV2540). Individuals with significantly immunocompromising conditions (ie, any condition or therapy that causes clinically significant immunosuppression) or with a history of myocarditis or pericarditis were excluded. Additionally, individuals of child-bearing potential were excluded by urine pregnancy test at baseline.

Neutralizing antibody (nAb) titers, anti-spike immunoglobulin G (IgG) antibody levels, and human angiotensin-converting enzyme 2 (hACE2) receptor binding inhibition titers were tested at baseline (Day 0, pre-dose) and Day 28. Safety was evaluated via adverse events of reactogenicity collected via memory aid for 7 days after vaccine administration, as well as vaccine-related medically attended adverse events (MAAEs), all serious adverse events (SAEs), and adverse events of special interest (AESIs), collected through the 6-month follow-up of the study.

### Immunogenicity testing

Testing of nAb titers using a validated pseudovirus-based neutralizing assay^18^ and of IgG antibody levels measured by validated anti-spike IgG enzyme-linked immunosorbent assay (ELISA),^19^ as well as of hACE2 receptor binding inhibition titers^20^ by the Clinical Immunology Laboratory of Novavax, Inc., Gaithersburg, MD, were performed on serum samples collected at the time of enrollment and Day 28 post vaccination.^17, 18^ The absence of previous or current SARS-CoV-2 infection was determined by non-reactive anti-nucleoprotein (anti-NP) antibodies and negative SARS-CoV-2 nasal PCR performed at baseline by the University of Washington Virology Laboratory, Seattle, WA.^3^ Validated immunogenicity assays^19^ were performed for the ancestral Wuhan antigen and for the Omicron BA.5 antigen. Fit-for-purpose pseudovirus-based neutralization assays were used to test for cross-reactivity against the more recent JN.1 and BA.2.86 lineages (using the same assay methodology as previously published^18^). Antibodies to Omicron BA.5 had not been measured in sera from Study 2019nCov-307; thus, no two-dose comparison is available for the variant nAb titers.

### Statistical analyses

The analysis populations for the study included the Safety population and the Per Protocol Immunogenicity 1 and 2 populations. These populations are defined as follows:

*Safety Analysis Set* – all study participants who provided consent were enrolled, received their dose of study vaccine, and provided safety data in follow-up.

*Per Protocol Analysis Set 1 (PP-IMM1)* – all study participants who received their assigned study vaccine according to the protocol with no receipt of another COVID-19 vaccine (authorized or approved) after Day 0 vaccination, who provided serology results for baseline and Day 28, with availability after vaccination, who were PCR negative at baseline for SARS-CoV-2, who did not report SARS-CoV-2 infection through the Day-28 blood draw, and who had no major protocol violations that were considered clinically relevant to impact the immunogenicity response. Exclusionary violations were determined by Novavax prior to database lock. A negative anti-NP result at baseline was not required. Participants with missing anti-NP results at baseline were included.

*Per Protocol Analysis Set 2 (PP-IMM2)* – all study participants who were defined similarly to Per Protocol Analysis Set 1 but who were additionally required to have a negative baseline anti-NP result.

Noninferiority of neutralizing antibody titers induced by two doses was based on the ratio of geometric mean titers (GMT) and the difference in proportion of participants with a seroresponse (also noted as seroconversion rate [SCR]) to vaccination (4-fold rise at Day 28 over baseline). The ratio comparing nAb GMT 28 days after the current study dose (NVX-CoV2373 in Study 2019nCoV-312) versus nAb GMT 28 days after the previous study dose (NVX-CoV2373 in Study 2019nCoV-307) and the 95% CIs, was calculated using paired *t*-test distribution.

The CIs of the difference in proportion of participants with SCR between 28 days after the current study dose (NVX-CoV2373 in Study 2019nCoV-312) and 28 days after the previous study dose (NVX-CoV2373 in Study 2019nCoV-307), both relative to the date of the dose in Study 2019nCoV-307, was calculated using the method of Tango (details in Statistical Analysis Plan, Supplemental Data). The criteria for demonstration of non-inferiority of the current study dose to the previous study dose required a lower bound (LB) of the 95% CIs for the ratio of nAb GMTs to be >0.67 and LB of the 95% CIs for differences in SCRs to be >−10%. If non-inferiority were demonstrated, testing for superiority was planned and was defined as LB of the 95% CIs for the ratio of nAb GMTs >1.0.

## RESULTS

### Study population

The ancestral strain vaccine group enrolled 104 participants with a median age of 40 years (range 19–50) who were approximately 61% female, 12% Hispanic, and 22% Black/African American. The Omicron BA.5 vaccine group, enrolled approximately 2 months later, was of the same age but with a slightly higher representation of males, Hispanics, and Black/African Americans (**Table 1**). Both groups had similar histories of prior COVID-19 vaccines, with >90% having received only homologous mRNA regimens prior to receipt of ancestral strain vaccine in Study 2019nCoV-307. Very few participants in either group reported having previously experienced symptomatic COVID-19, but over 70% were seropositive for anti-NP at baseline, indicating prior exposure. The disposition of study participants is outlined in **Fig 1**.

**FIG 1.**
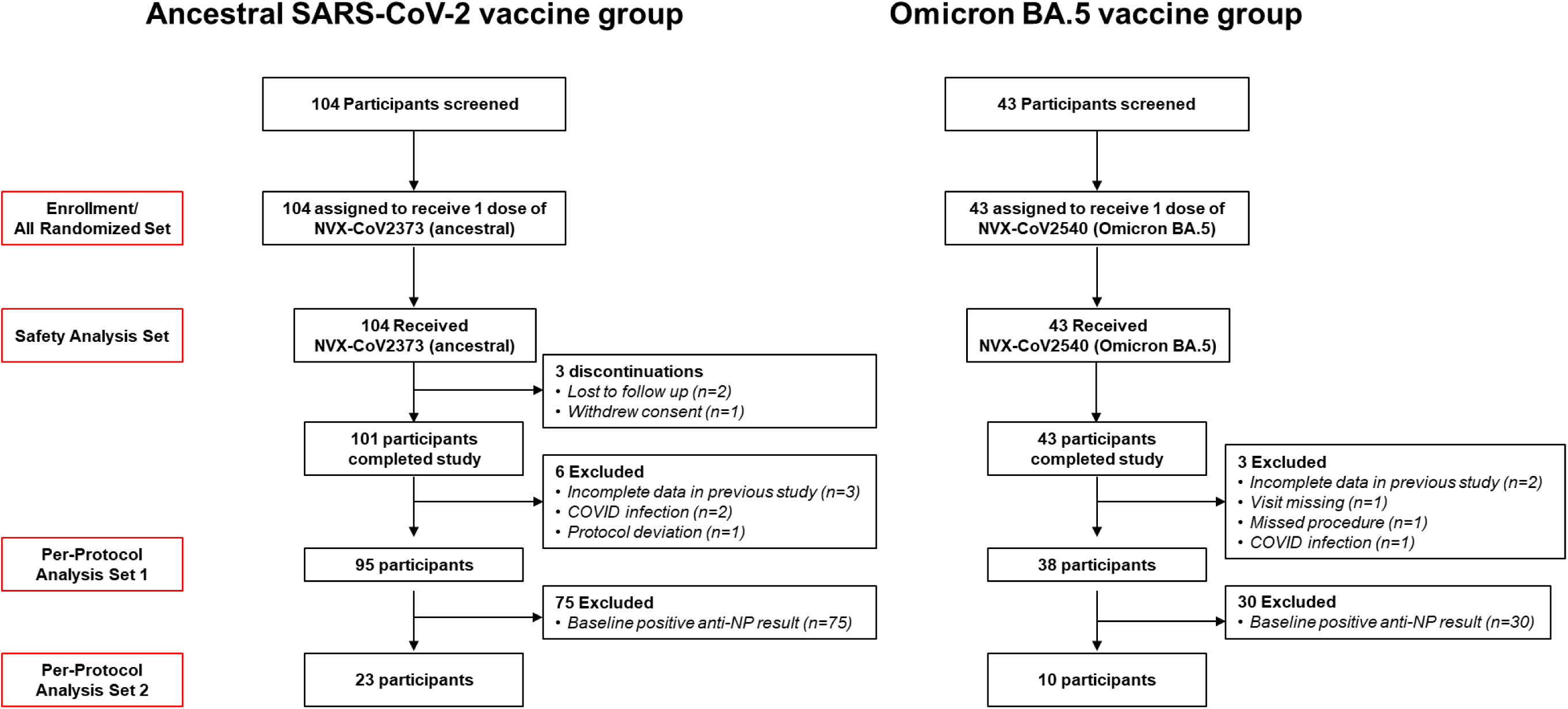
CONSORT diagram. NP, nucleocapsid.

**Table 1.**
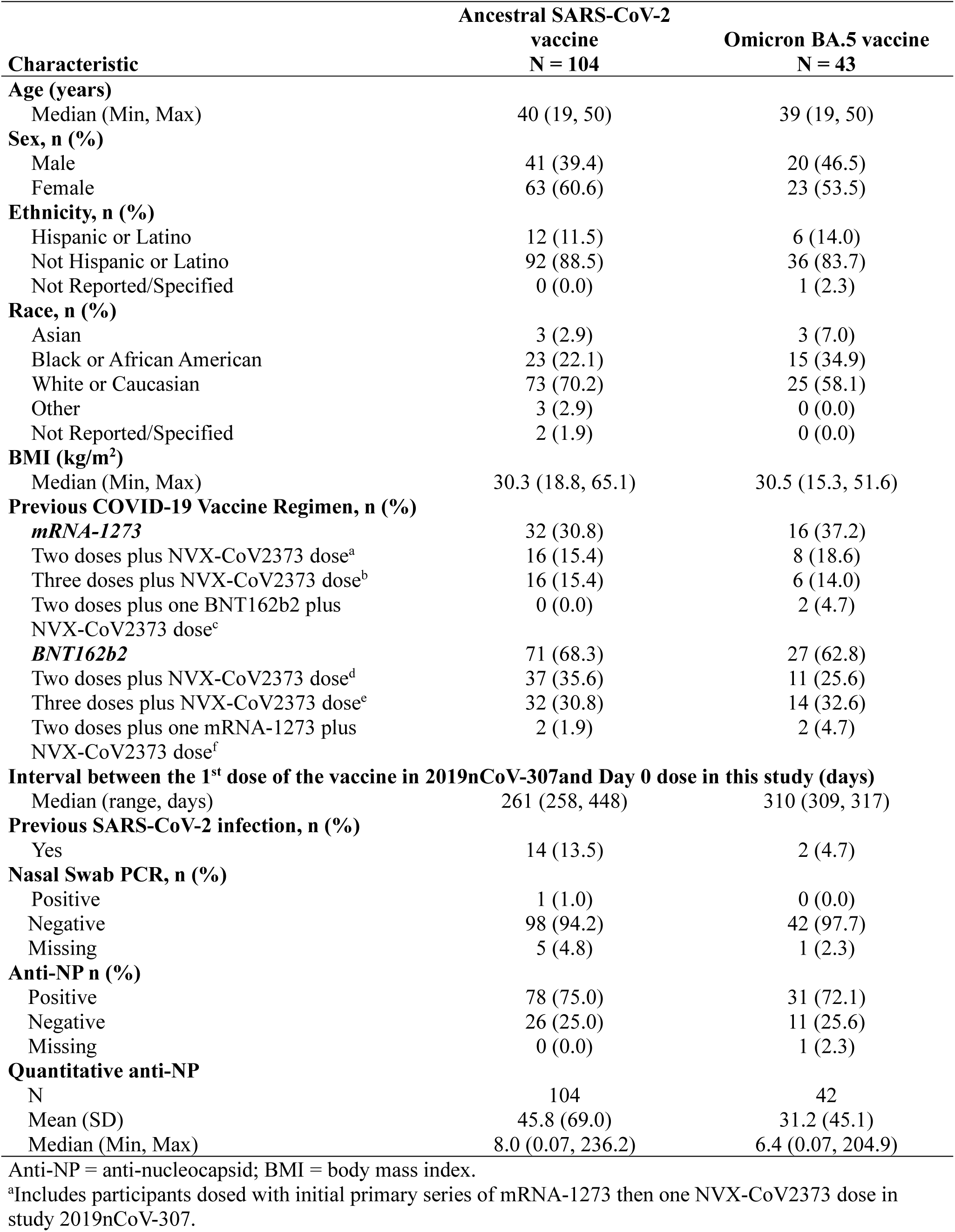
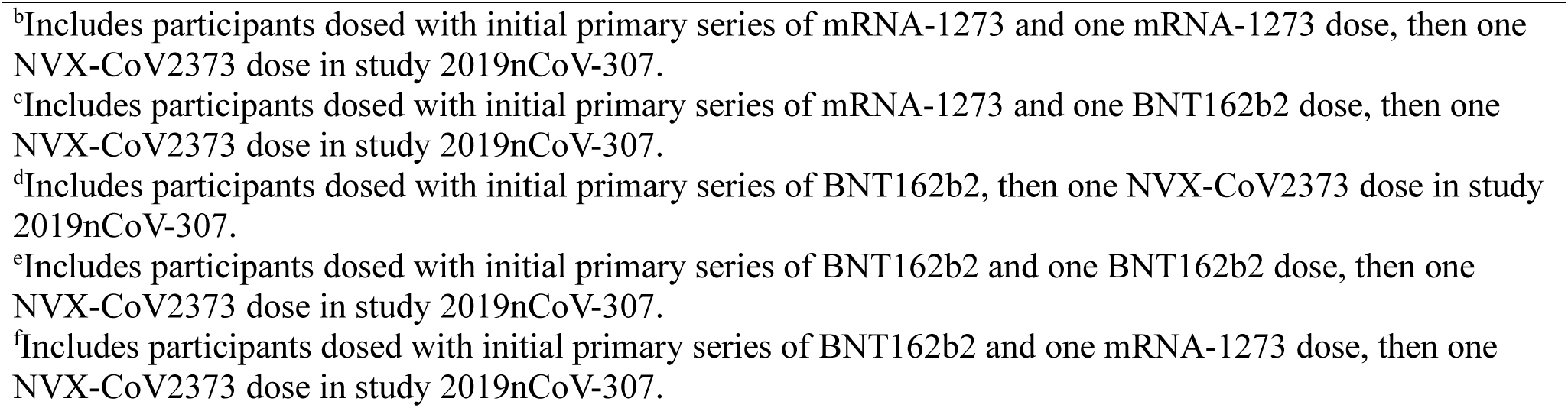
Demographics and baseline characteristics (Safety analysis set).

| Characteristic | Ancestral SARS-CoV-2<br>vaccine<br>N = 104 | Omicron BA.5 vaccine<br>N = 43 |
| --- | --- | --- |
| **Age (years)** |  |  |
| Median (Min, Max) | 40 (19, 50) | 39 (19, 50) |
| **Sex, n (%)** |  |  |
| Male | 41 (39.4) | 20 (46.5) |
| Female | 63 (60.6) | 23 (53.5) |
| **Ethnicity, n (%)** |  |  |
| Hispanic or Latino | 12 (11.5) | 6 (14.0) |
| Not Hispanic or Latino | 92 (88.5) | 36 (83.7) |
| Not Reported/Specified | 0 (0.0) | 1 (2.3) |
| **Race, n (%)** |  |  |
| Asian | 3 (2.9) | 3 (7.0) |
| Black or African American | 23 (22.1) | 15 (34.9) |
| White or Caucasian | 73 (70.2) | 25 (58.1) |
| Other | 3 (2.9) | 0 (0.0) |
| Not Reported/Specified | 2 (1.9) | 0 (0.0) |
| **BMI (kg/m<sup>2</sup>)** |  |  |
| Median (Min, Max) | 30.3 (18.8, 65.1) | 30.5 (15.3, 51.6) |
| **Previous COVID-19 Vaccine Regimen, n (%)** |  |  |
| **mRNA-1273** | 32 (30.8) | 16 (37.2) |
| Two doses plus NVX-CoV2373 dose<sup>a</sup> | 16 (15.4) | 8 (18.6) |
| Three doses plus NVX-CoV2373 dose<sup>b</sup> | 16 (15.4) | 6 (14.0) |
| Two doses plus one BNT162b2 plus NVX-CoV2373 dose<sup>c</sup> | 0 (0.0) | 2 (4.7) |
| **BNT162b2** | 71 (68.3) | 27 (62.8) |
| Two doses plus NVX-CoV2373 dose<sup>d</sup> | 37 (35.6) | 11 (25.6) |
| Three doses plus NVX-CoV2373 dose<sup>e</sup> | 32 (30.8) | 14 (32.6) |
| Two doses plus one mRNA-1273 plus NVX-CoV2373 dose<sup>f</sup> | 2 (1.9) | 2 (4.7) |
| **Interval between the 1<sup>st</sup> dose of the vaccine in 2019nCoV-307 and Day 0 dose in this study (days)** |  |  |
| Median (range, days) | 261 (258, 448) | 310 (309, 317) |
| **Previous SARS-CoV-2 infection, n (%)** |  |  |
| Yes | 14 (13.5) | 2 (4.7) |
| **Nasal Swab PCR, n (%)** |  |  |
| Positive | 1 (1.0) | 0 (0.0) |
| Negative | 98 (94.2) | 42 (97.7) |
| Missing | 5 (4.8) | 1 (2.3) |
| **Anti-NP n (%)** |  |  |
| Positive | 78 (75.0) | 31 (72.1) |
| Negative | 26 (25.0) | 11 (25.6) |
| Missing | 0 (0.0) | 1 (2.3) |
| **Quantitative anti-NP** |  |  |
| N | 104 | 42 |
| Mean (SD) | 45.8 (69.0) | 31.2 (45.1) |
| Median (Min, Max) | 8.0 (0.07, 236.2) | 6.4 (0.07, 204.9) |

Anti-NP = anti-nucleocapsid; BMI = body mass index.
<sup>a</sup>Includes participants dosed with initial primary series of mRNA-1273 then one NVX-CoV2373 dose in study 2019nCoV-307.
<sup>b</sup>Includes participants dosed with initial primary series of mRNA-1273 and one mRNA-1273 dose, then one NVX-CoV2373 dose in study 2019nCoV-307.
<sup>c</sup>Includes participants dosed with initial primary series of mRNA-1273 and one BNT162b2 dose, then one NVX-CoV2373 dose in study 2019nCoV-307.
<sup>d</sup>Includes participants dosed with initial primary series of BNT162b2, then one NVX-CoV2373 dose in study 2019nCoV-307.
<sup>e</sup>Includes participants dosed with initial primary series of BNT162b2 and one BNT162b2 dose, then one NVX-CoV2373 dose in study 2019nCoV-307.
<sup>f</sup>Includes participants dosed with initial primary series of BNT162b2 and one mRNA-1273 dose, then one NVX-CoV2373 dose in study 2019nCoV-307.

### Immunogenicity results

#### Neutralizing antibodies

##### NVX-CoV2373 group – Ancestral Wuhan-based study vaccine

The primary immunogenicity objective to demonstrate noninferiority of this study dose (ancestral) versus previous study dose, measured by anti-Wuhan nAb at Day 28, was achieved. As described in the Supplemental Methods, similar validated pseudovirus-based neutralization assays from two different laboratories were used in the current and previous study; to allow comparisons across studies, GMTs in Study 2019nCoV-307 were converted from ID_50_ to IU/mL. This analysis compared GMTs 28 days after the dose in this study (393.2 IU/mL (95% CI: 318.0–468.2) versus 28 days after the dose in the previous study (396.6 IU/mL (95% CI: 328.7–478.6) (**Fig 2, A**). The ratio (calculated using paired *t*-test to account for the within-subject variability) of nAb GMTs at Day 28 between this study and previous study doses was 1.0 (95% CI: 0.8–1.2), and the LB was higher than the prespecified criterion of 0.67 (**Fig 2, B**).

**FIG 2.**
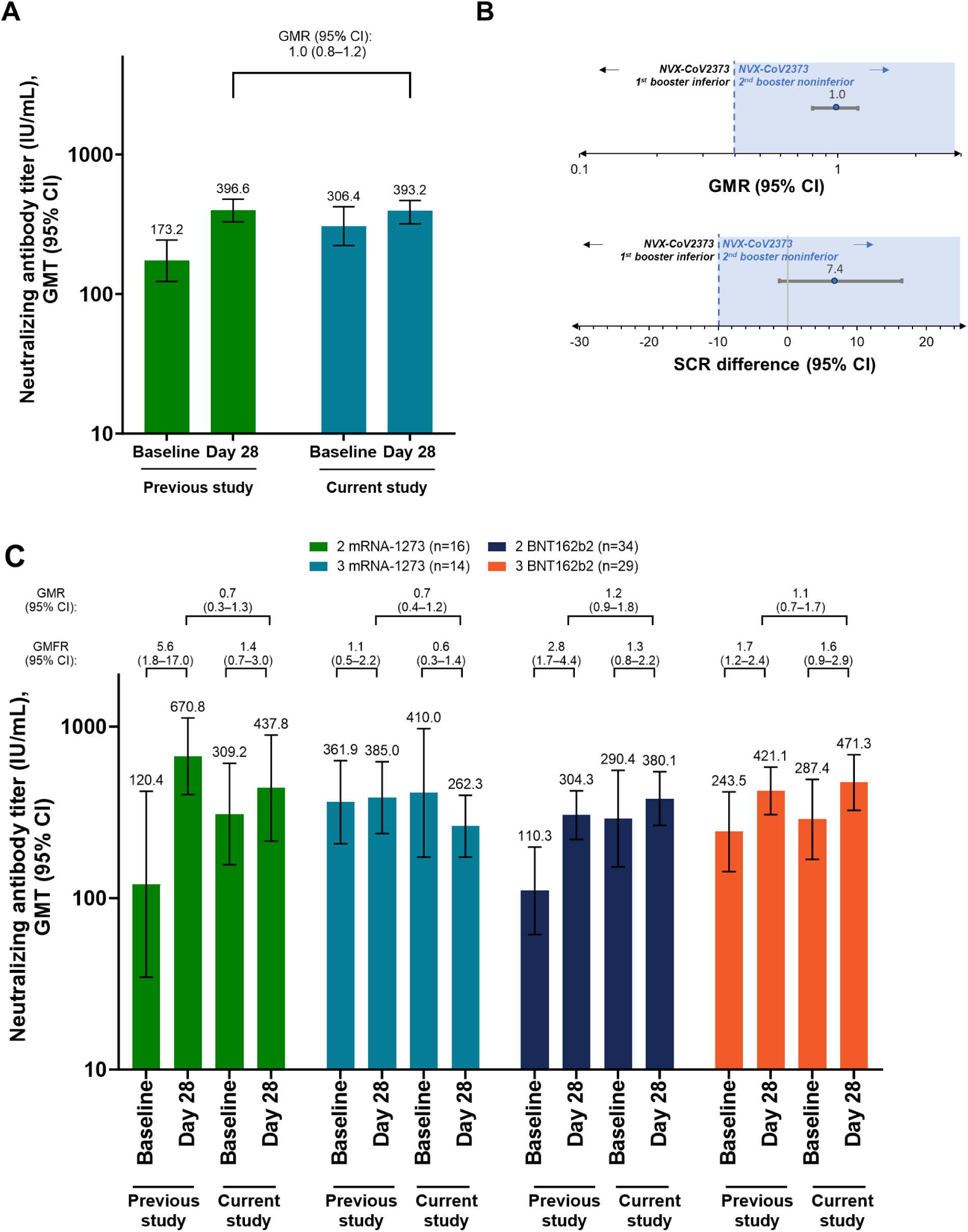
Pseudovirus-based nAb against the ancestral strain after ancestral SARS-CoV-2 vaccine. Data are shown for (A) nAbs in the PP1 population and (B) the corresponding analyses of noninferiority for GMR and SCR difference, and (C) assessed by prior vaccination. Participants who received an initial primary series of BNT162b2, 1 mRNA-1273 dose, and 1 prior NVX-CoV2373 dose before the previous study are not shown in part C because of small sample sizes (n = 2). There were no participants who received an initial primary series of mRNA-1273, 1 BNT162b2 dose, and 1 prior NVX-CoV2373 dose before the previous study.

Similarly, the secondary immunogenicity noninferiority criterion of difference in seroresponse rates following doses in this study (22.1% [95% CI: 14.2–31.8]) versus the previous study (25.3% [95% CI: 16.9–35.2]) was 7.4% (−1.2 to 16.5), also meeting noninferiority (LB >−10%) (**Fig 2, B**).

Quantitation of nAb titers after the current study dose versus the previous study according to prior mRNA vaccination history (ie, whether two or three doses of either mRNA vaccine [mRNA-1273 or BNT162b2]) showed no clinically meaningful overall difference across the groups (**Fig 2, C**).

Encouragingly, nAb titers induced by a dose of ancestral Wuhan strain vaccine in the current study that cross-reacted with Omicron BA.5 showed substantial rise in levels (329.0 ID_50_ [95% CI: 241.0–449.1]) prior to and 28 days following (911.5 [711.3–1168.2]) the current study dose, suggesting a likely degree of cross-protection against emerging variant strains (**Fig 3, A**). The magnitude of Omicron BA.5 cross-reactive nAb rise induced by the ancestral Wuhan vaccine was even higher in participants in the current study without prior infection (anti-NP seronegative) at baseline (84.5 ID_50_ [95% CI: 55.8–128.0] and 28 days after dose (451.3 ID_50_ [95% CI: 242.1–841.3]) (**Fig 3, B**).

**FIG 3.**
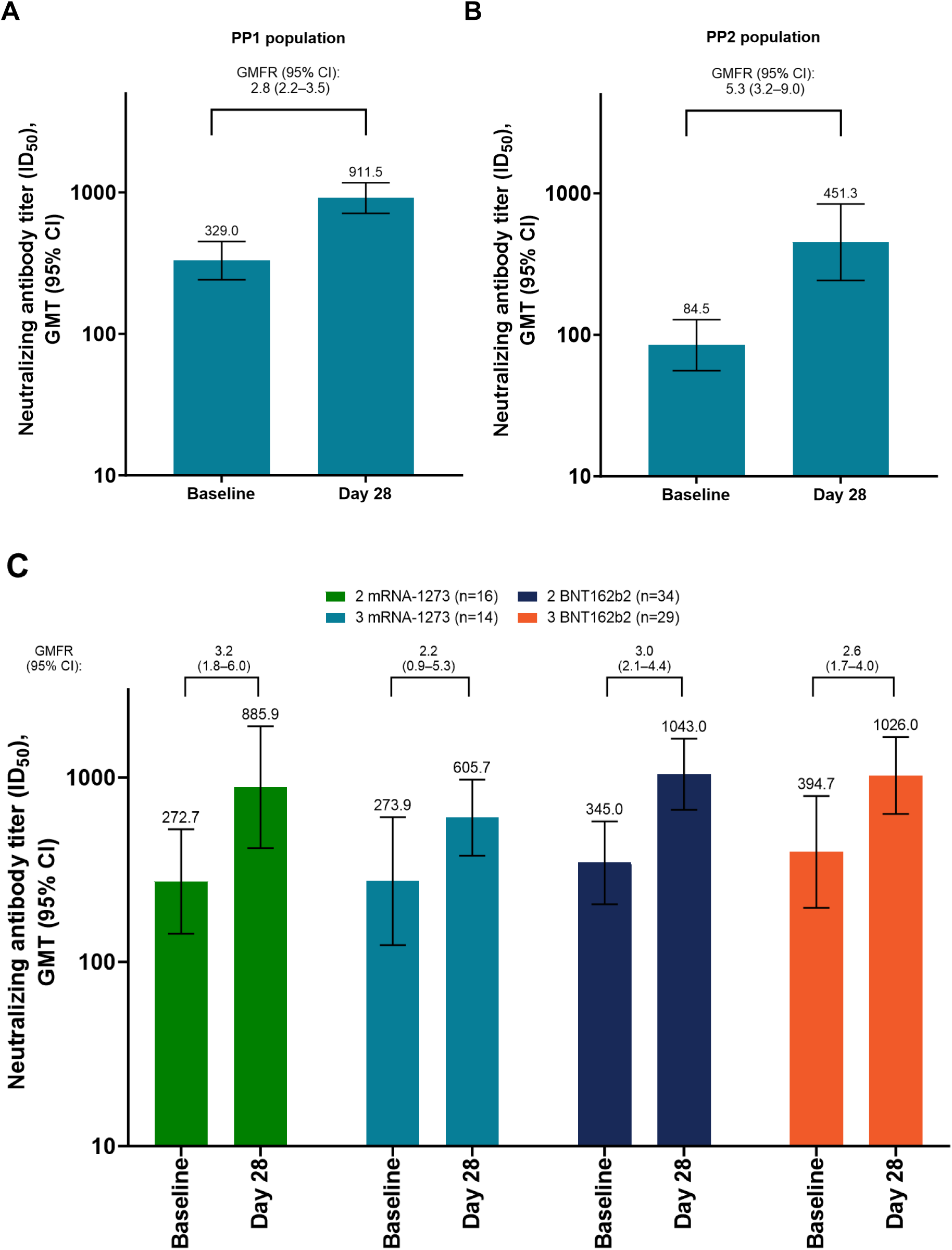
Pseudovirus-based nAb against SARS-CoV-2 Omicron BA.5 after ancestral SARS-CoV-2 vaccine. (A) PP1 population, (B) PP2 population, (C) PP1 population, assessed by prior vaccination. Participants who received an initial primary series of BNT162b2, 1 mRNA-1273 dose, and 1 prior NVX-CoV2373 dose before the previous study are not shown in part C because of small sample sizes (n = 2). There were no participants who received an initial primary series of mRNA-1273, 1 BNT162b2 dose, and 1 prior NVX-CoV2373 dose before the previous study.

Cross-reactive nAbs to BA.5 after a second ancestral Wuhan vaccine also showed no meaningful difference relative to the prior mRNA vaccination history **(Fig 3, C)**.

##### NVX-CoV2540 group – Omicron BA.5 variant study vaccine

In all participants who received the Omicron BA.5 vaccine, nAb GMTs (ID_50_ [95% CI]) against the SARS-CoV-2 Omicron BA.5 variant increased from 305.8 (170.5, 548.4) at baseline to 2213 (1368.0, 3580.1) 28 days after the current study dose, which shows a substantial geometric mean fold rise (GMFR) of 7.2 (95% CI: 4.9–10.7]) **(Fig 4, A)**. Among the participants anti-NP–negative at the time of the current study dose, Omicron BA.5 nAbs induced by the Omicron BA.5 vaccine were similarly robust from 91.7 (28.4, 295.8) to 1809 (573.0, 5710.6), pre- and 28 days post-dose, respectively (**Fig 4, B**).

**FIG 4.**
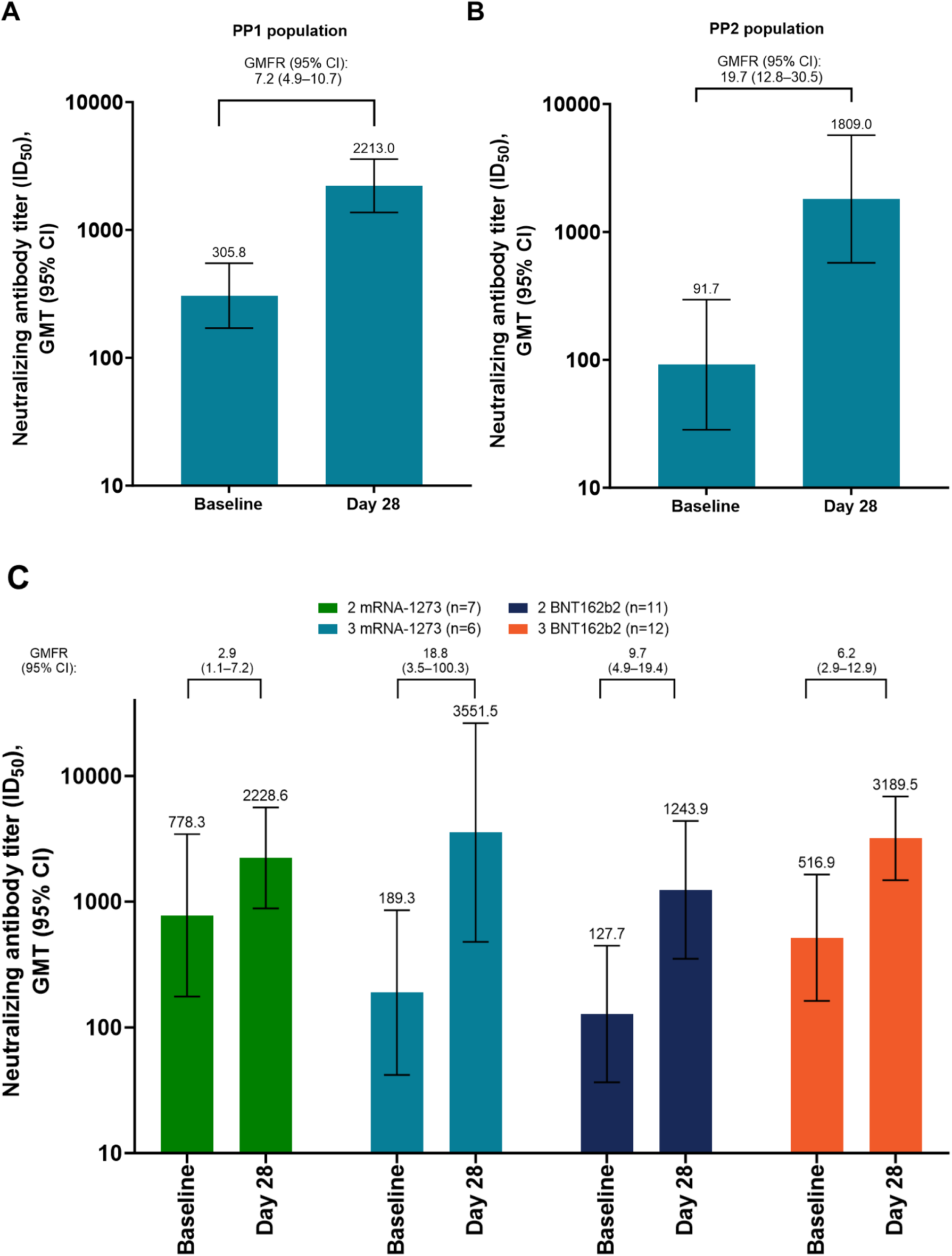
Pseudovirus-based nAb against SARS-CoV-2 Omicron BA.5 after Omicron BA.5 vaccine. (A) PP1 population, (B) PP2 population, (C) PP1 population, assessed by prior vaccination. Participants who received an initial primary series of BNT162b2, 1 mRNA-1273 dose, and 1 prior NVX-CoV2373 dose before the previous study are not shown in part C because of small sample sizes (n = 2). There were no participants who received an initial primary series of mRNA-1273, 1 BNT162b2 dose, and 1 prior NVX-CoV2373 dose before the previous study.

There was no apparent difference in responses based on prior mRNA vaccination history (**Fig 4, C**). The suggestion of a more robust BA.5 nAb response to the Omicron BA.5 vaccine among participants with a prior history of three doses of mRNA vaccine might be attributable to the very small sample sizes, as confidence limits were very wide.

Conversely, a similar substantial cross-reacting response to the ancestral Wuhan strain was observed among participants whose current study dose was the BA.5 study vaccine: 420.7 IU/mL (95% CI: 282.5– 626.6) and 835.0 IU/mL (95% CI: 597.1–1167.6) pre- and 28 days post-dose, respectively (**Fig 5, A**).

**FIG 5.**
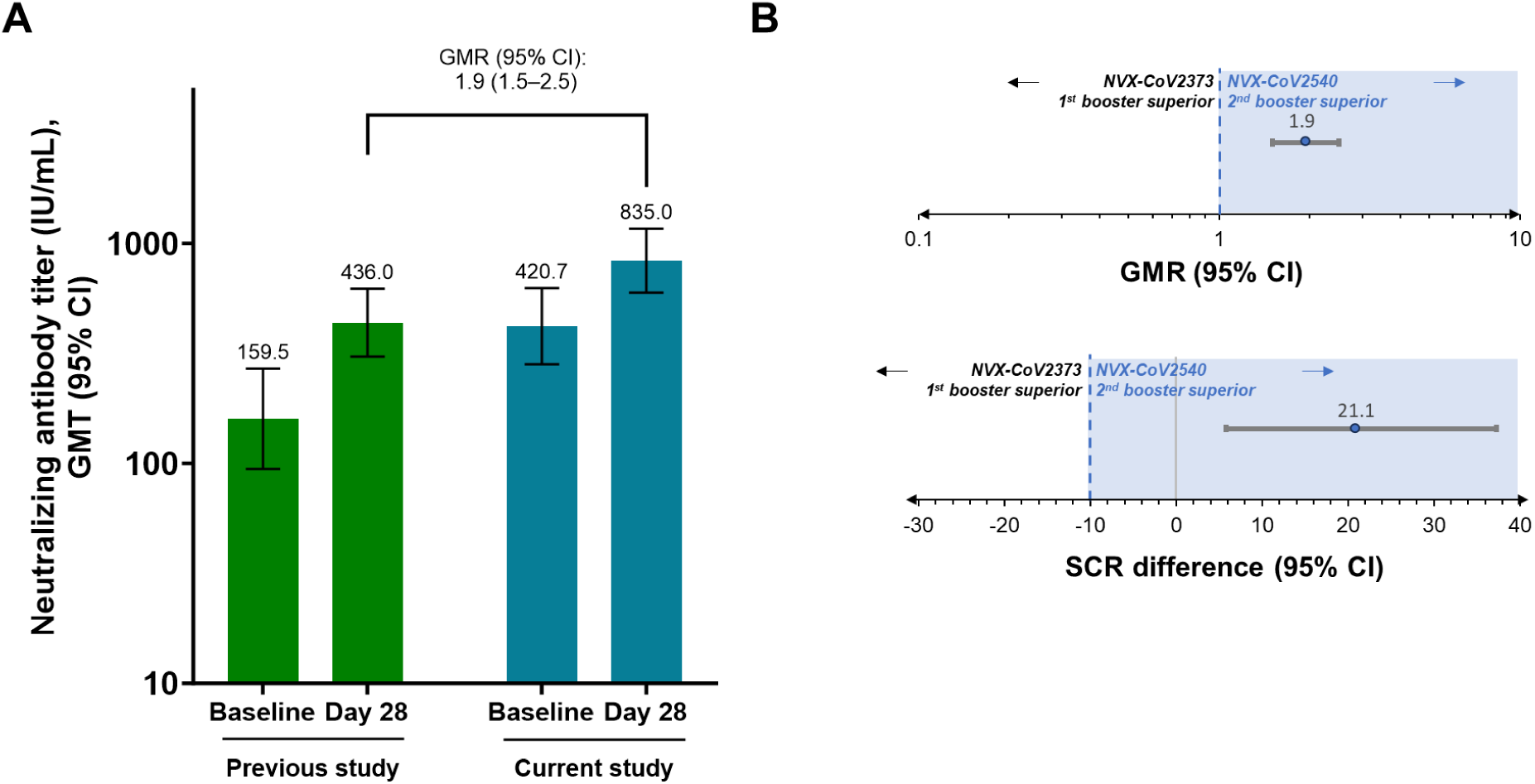
Pseudovirus-based nAb against the SARS-CoV-2 ancestral strain after a dose of Omicron BA.5 vaccine. Data are shown for (A) nAbs in the PP1 population and (B) the corresponding analyses of superiority for GMR and SCR difference.

Neutralizing antibody titers to the Wuhan strain induced by a dose of Omicron BA.5 in the current study were superior to those induced by a dose of ancestral Wuhan vaccine strain in the current study: GMFR of 1.9 (95% CI: 1.5–2.5) and a difference in SCRs of 21.1% (95% CI: 5.8–37.3) (**Fig 5, B**). Among the anti-NP–negative participants at the time of the Omicron BA.5 dose in the current study, Wuhan nAbs responses were similarly robust: 571.2 IU/mL (95% CI: 219.1–1488.9) at baseline and 931.4 IU/mL (95% CI: 405.3–2140.2) 28 days post-dose in the current study, which results in a GMFR of 7.4 (95% CI: 1.7– 32.3).

##### Cross-reactivity to newer SARS-CoV-2 variants

Both the ancestral and Omicron BA.5 variant strain vaccines elicited robust nAbs against other more recent Omicron variants, including JN.1 and BA.2.86 (**Fig S1**). When comparing Day 28 nAbs with the baseline levels before the current study dose, the Omicron BA5 vaccine tended to result in greater rises in antibody titers to Omicron variants than the ancestral vaccine.

##### Anti-spike IgG antibodies and hACE2 receptor binding inhibition titers

Anti-spike IgG levels measured by ELISA and serum hACE2 receptor binding inhibition titers were measured at the same time points and showed comparable overall results (**Fig S2-S5**).

### Safety results

### Reactogenicity

Overall, the incidence of reported local reactions was similar between the ancestral (77.9%) and Omicron BA.5 (74.4%) vaccine groups (**Fig 6**). The most frequently reported reactions were injection-site pain and tenderness, which were similar between groups. Local reactions of greater severity than mild or moderate were rare. Systemic reactions were also similar between the two groups (ancestral: 66.3% vs Omicron BA.5: 55.8%). The most commonly reported systemic reactions were fatigue (ancestral: 47.1% vs Omicron BA.5: 25.6%) and muscle pain (ancestral: 41.3% vs Omicron BA.5: 44.2%) (**Fig 6**).

**FIG 6.**
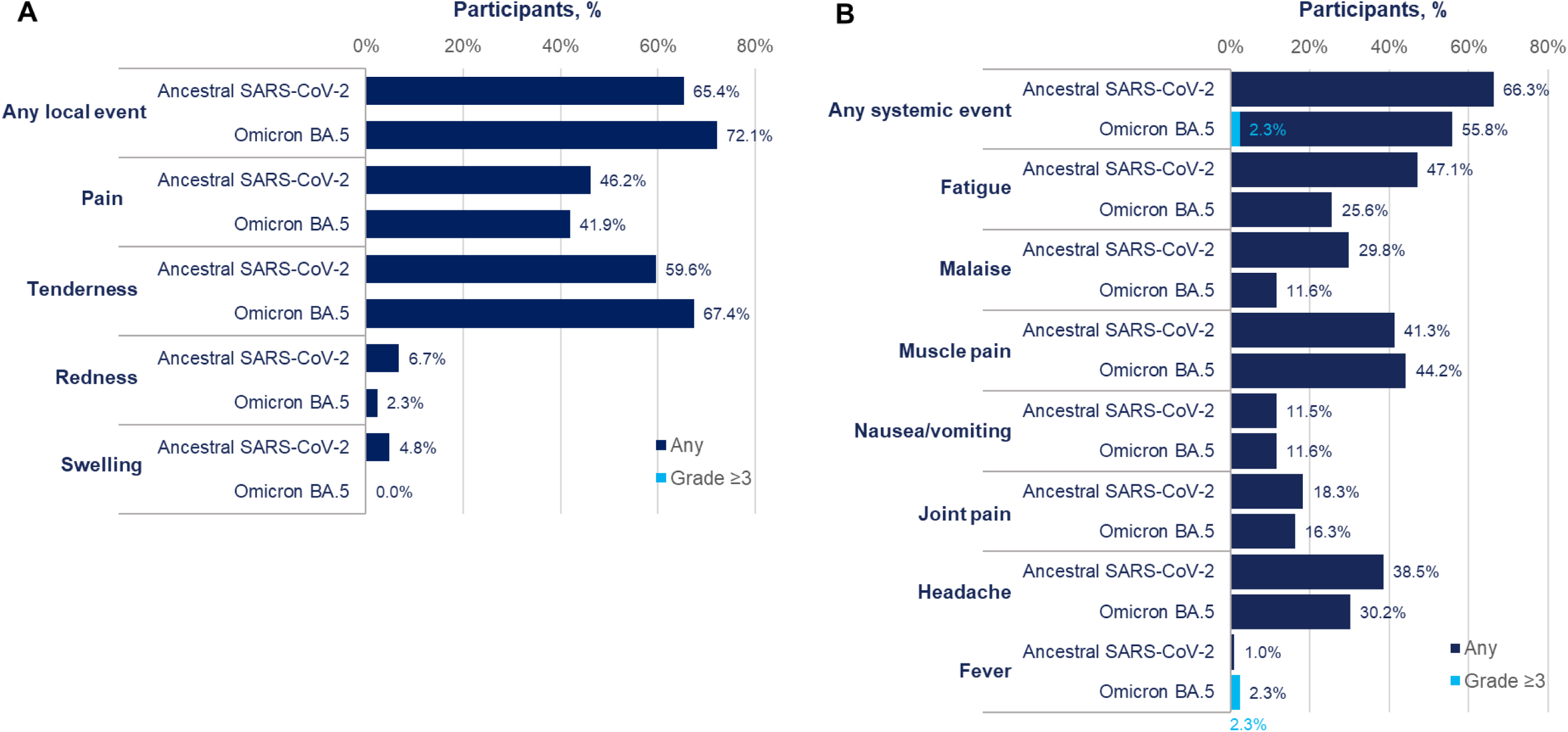
(A) Local and (B) systemic reactogenicity after vaccination in the current study with either the ancestral SARS-CoV-2–based vaccine or the Omicron BA.5–based vaccine

### Unsolicited Adverse Events

Unsolicited adverse events were reported very infrequently and only a single report of axillary lymphadenopathy, likely attributed to reactogenicity, was considered related to the study vaccines **(Table 2).**

**Table 2.**
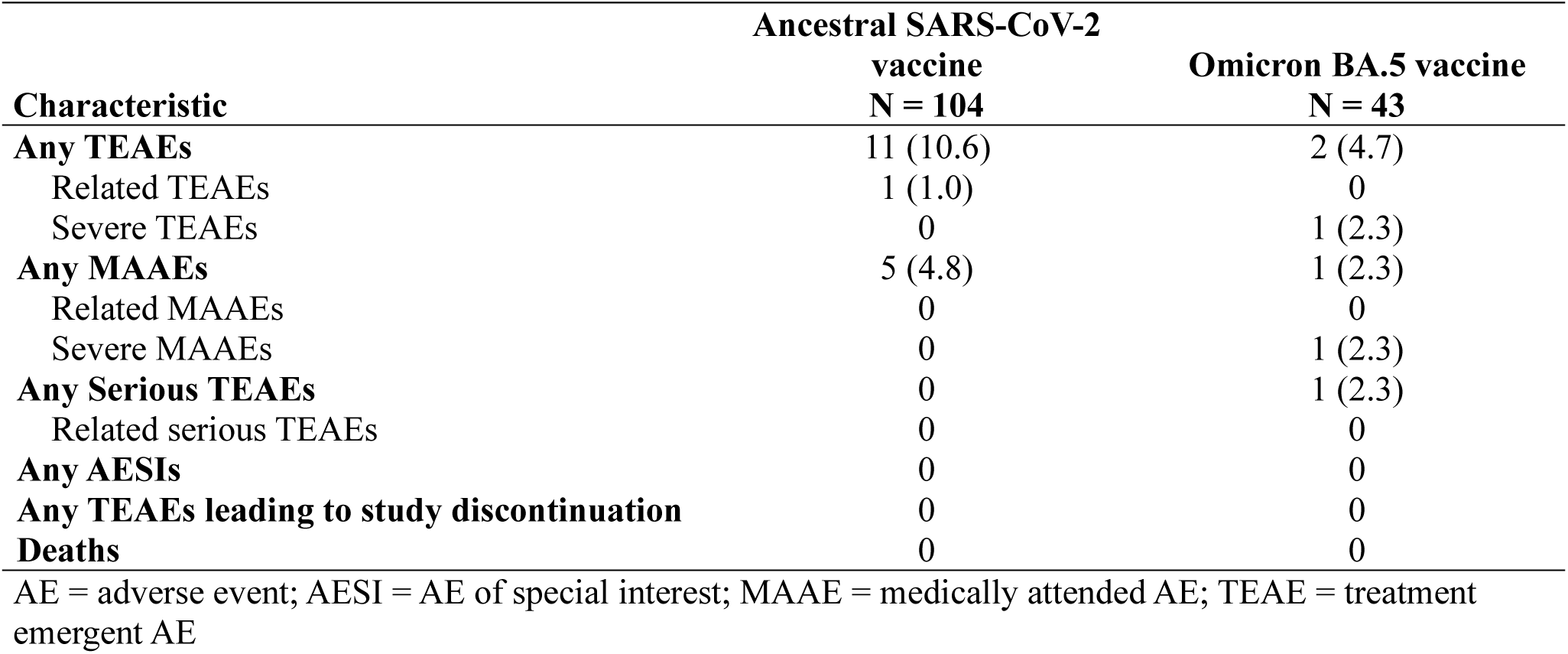
Unsolicited AEs through the end of the study (Safety analysis set).

| **Characteristic** | **Ancestral SARS-CoV-2** | **Omicron BA.5 vaccine** |
| --- | --- | --- |
|  | **vaccine**<br>**N = 104** | **N = 43** |
| **Any TEAEs** | 11 (10.6) | 2 (4.7) |
| Related TEAEs | 1 (1.0) | 0 |
| Severe TEAEs | 0 | 1 (2.3) |
| **Any MAAEs** | 5 (4.8) | 1 (2.3) |
| Related MAAEs | 0 | 0 |
| Severe MAAEs | 0 | 1 (2.3) |
| **Any Serious TEAEs** | 0 | 1 (2.3) |
| Related serious TEAEs | 0 | 0 |
| **Any AESIs** | 0 | 0 |
| **Any TEAEs leading to study discontinuation** | 0 | 0 |
| **Deaths** | 0 | 0 |

AE = adverse event; AESI = AE of special interest; MAAE = medically attended AE; TEAE = treatment emergent AE

## DISCUSSION AND CONCLUSIONS

With the evolution of the SARS-CoV-2 pandemic into an endemic situation, periodic outbreaks can occur due to immune-evasive mutations^21–23^ and subsequent antigenic drift, necessitating repeated immunization with updated vaccines based on evolving variants. Due to the availability of different vaccine types with unique mechanisms of action, it is desirable to determine the safety and immunogenicity of heterologous versus homologous series of vaccinations.

The adjuvanted, rS ancestral SARS-CoV-2 strain vaccine (NVX-CoV2373) demonstrated a high degree of protective efficacy following a two-dose primary series in SARS-CoV-2 infection-naïve individuals.^3^ In addition, the protein-based vaccine has been evaluated for immunogenicity and safety as an additional single heterologous dose following the initial series of mRNA vaccines.^3^ This study was designed to enroll individuals from a previously conducted study (Study 2019nCoV-307) and to assess the safety and immunogenicity of an additional dose of the adjuvanted rS vaccine based on the ancestral Wuhan strain as well as an updated vaccine formulation based on the Omicron BA.5 variant among participants who had received a two-dose primary series of mRNA vaccine with/without an additional homologous or heterologous mRNA dose prior to the 2019nCoV-307 study participation.

All the participants enrolled in the current study had previously received two or three doses of approved mRNA vaccines and a single dose of ancestral Wuhan-adjuvanted rS protein vaccine. At least 180 days after the most recent vaccine dose, another dose of ancestral vaccine elicited robust immune responses to the ancestral strain and a robust cross-reactive response to the Omicron BA.5 variant strain. Similarly, an updated Omicron BA.5 vaccine induced robust responses to Omicron BA.5 antigen and displayed cross-reactivity to the ancestral Wuhan virus strain. The magnitude of these responses was not meaningfully different among participants based on their prior vaccination history, regardless of whether two or three prior doses of mRNA vaccine had been administered and irrespective of the specific mRNA vaccine type (Moderna or Pfizer) received previously. This finding might be due to an immune response plateau above which neutralizing (and likely other) antibodies appear not to rise linearly based upon modeling.^24^

The safety profile defined by 7 days of solicited reactions and 6 months of unsolicited adverse events following either vaccine strain proved to be clinically acceptable and comparable to earlier studies.^3, 6, 17^

The major limitation of this study, especially of the Omicron BA.5 group analysis, is the small sample size overall and in subsets defined by prior vaccination history. While there is consistency in the findings, the groups are too small to draw statistical conclusions. Omicron BA.5 nAbs were not measured in sera from Study 2019nCov-307; thus, no comparison of the current study dose versus the previous study dose is available for the variant titers.

The responses to the Omicron BA.5 variant–based vaccine, especially cross-reactive antibodies to the newer variants, are relevant as the circulating strains of SARS-CoV-2 evolve and new variants completely replace older variants. Accordingly, while these analyses provide encouraging findings suggesting that some degree of cross-protection may be expected from vaccines based on earlier variants, it is likely that vaccines representing more recent strains might be required on a periodic basis. It is encouraging that both the ancestral and Omicron BA.5 vaccines elicited nAb titers that cross-reacted with more recent JN.1 and BA.2.8.6 variants.

Results from this study suggest that multiple doses of the adjuvanted rS protein–based vaccine can be administered safely in sequential heterologous series of vaccines and can produce robust immune responses regardless of prior COVID-19 vaccine history with respect to vaccine platform/type or number of doses.

## Data Availability

Study information is available online at links provided. Requests submitted to the corresponding author will be considered.

https://www.clinicaltrials.gov/study/NCT05875701?term=2019nCoV-312&rank=1

https://www.clinicaltrials.gov/study/NCT05463068?term=NCT05463068&rank=1

## Acknowledgements

We thank all of the study participants who volunteered for this study. We also thank Miranda R. Cai, PhD, and Kesava Vajjala, of Novavax, Inc. for statistical support. Medical writing, editing, and formatting support, under the direction of the authors, was provided by Kelly M Cameron, PhD, CMPP, and Ebenezer M. Awuah-Yeboah, BS, of Ashfield MedComms, an Inizio company, and was funded by Novavax, Inc. Medical writing, editing, and formatting support was also provided by Anthony M. Marchese, PhD, and Anar Murphy, PhD, CMPP, of Novavax, Inc.

## Contributions

Study design and conception: RMM, LMD

Data extraction and analysis: JMA, PB, MRC, GC

Interpretation of data: JMA, PB, MRC, GC, RK, SCC, MZ, ZC, ME, JSP, RMM, and LMD

Data verification: MRC, GC

Project management: ME, LMD Writing: first draft - LMD

Writing: reviewing and editing: JMA, PB, MRC, GC, RK, SCC, MZ, ZC, ME, JSP, RMM, and LMD

Statistical analysis: MRC, GC

All authors reviewed and provided final approval for submission of this manuscript.

## Data sharing statement

Study information is available online at https://www.clinicaltrials.gov/study/NCT05875701?term=2019nCoV-312&rank=1 for the current study and https://www.clinicaltrials.gov/study/NCT05463068?term=NCT05463068&rank=1 for the previous study. Requests submitted to the corresponding author will be considered.

## References

1. COVID Data Tracker. 2024.] Available at: https://covid.cdc.gov/covid-data-tracker/#datatracker-home.

2. WHO COVID-19 dashboard. 2024.] Available at: https://data.who.int/dashboards/covid19/cases.

3. Dunkle LM, Kotloff KL, Gay CL, Anez G, Adelglass JM, Barrat Hernandez AQ, et al. Efficacy and safety of NVX-CoV2373 in adults in the United States and Mexico. N Engl J Med 2022; 386:531–43.

4. El Sahly HM, Baden LR, Essink B, Doblecki-Lewis S, Martin JM, Anderson EJ, et al. Efficacy of the mRNA-1273 SARS-CoV-2 vaccine at completion of blinded phase. N Engl J Med 2021; 385:1774–85.

5. Falsey AR, Sobieszczyk ME, Hirsch I, Sproule S, Robb ML, Corey L, et al. Phase 3 safety and efficacy of AZD1222 (ChAdOx1 nCoV-19) covid-19 vaccine. N Engl J Med 2021; 385:2348–60.

6. Heath PT, Galiza EP, Baxter DN, Boffito M, Browne D, Burns F, et al. Safety and efficacy of NVX-CoV2373 covid-19 vaccine. N Engl J Med 2021; 385:1172–83.

7. Polack FP, Thomas SJ, Kitchin N, Absalon J, Gurtman A, Lockhart S, et al. Safety and efficacy of the BNT162b2 mRNA covid-19 vaccine. N Engl J Med 2020; 383:2603–15.

8. Sadoff J, Gray G, Vandebosch A, Cardenas V, Shukarev G, Grinsztejn B, et al. Safety and efficacy of single-dose Ad26.COV2.S vaccine against covid-19. N Engl J Med 2021; 384:2187-201.

9. EMA and ECDC statement on updating COVID-19 vaccines to target new SARS-CoV-2 virus variants. 2023.] Available at: https://www.ema.europa.eu/en/news/ema-and-ecdc-statement-updating-covid-19-vaccines-target-new-sars-cov-2-virus-variants#:~:text=In%20line%20with%20the%20outcome,other%20parts%20of%20the%20world.

10. Recommendations for the 2023-2024 formula of COVID-19 vaccines in the U.S.; 2023.] Available at: https://www.fda.gov/media/169591/download#:~:text=reviewed%20manufacturing%20timelines.-,For%20the%202023-2024%20Formula%20of%20COVID-19%20vaccines%20in,1.5.

11. Statement on the antigen composition of COVID-19 vaccines. 2023.] Available at: https://www.who.int/news/item/18-05-2023-statement-on-the-antigen-composition-of-covid-19-vaccines.

12. Updated COVID-19 vaccines for use in the United States beginning in fall 2024. 2024.] Available at: https://www.fda.gov/vaccines-blood-biologics/updated-covid-19-vaccines-use-united-states-beginning-fall-2024.

13. Baden LR, El Sahly HM, Essink B, Follmann D, Hachigian G, Strout C, et al. Long-term safety and effectiveness of mRNA-1273 vaccine in adults: COVE trial open-label and booster phases. Nat Commun 2024; 15:7469.

14. Moreira ED, Jr., Kitchin N, Xu X, Dychter SS, Lockhart S, Gurtman A, et al. Safety and efficacy of a third dose of BNT162b2 covid-19 vaccine. N Engl J Med 2022; 386:1910–21.

15. Novovax COVID-19 vaccine [package insert]. Gaithersburg, MD. Novavax, Inc. 2024.

16. Mbaeyi S, Oliver SE, Collins JP, Godfrey M, Goswami ND, Hadler SC, et al. The Advisory Committee on Immunization Practices’ interim recommendations for additional primary and booster doses of COVID-19 vaccines - United States, 2021. MMWR Morb Mortal Wkly Rep 2021; 70:1545-52.

17. Raiser F, Davis M, Adelglass J, Cai MR, Chau G, Cloney-Clark S, et al. Immunogenicity and safety of NVX-CoV2373 as a booster: a phase 3 randomized clinical trial in adults. Vaccine 2023; 41:5965–73.

18. Cai Z, Kalkeri R, Wang M, Haner B, Dent D, Osman B, et al. Validation of a pseudovirus neutralization assay for severe acute respiratory syndrome coronavirus 2: a high-throughput method for the evaluation of vaccine immunogenicity. Microorganisms 2024; 12.

19. Zhu M, Cloney-Clark S, Feng SL, Parekh A, Gorinson D, Silva D, et al. A severe acute respiratory syndrome coronavirus 2 anti-spike immunoglobulin G assay: a robust method for evaluation of vaccine immunogenicity using an established correlate of protection. Microorganisms 2023; 11.

20. Plested JS, Zhu M, Cloney-Clark S, Massuda E, Patel U, Klindworth A, et al. Severe acute respiratory syndrome coronavirus 2 receptor (human angiotensin-converting Enzyme 2) binding inhibition assay: a rapid, high-throughput assay useful for vaccine immunogenicity evaluation. Microorganisms 2023; 11.

21. Carabelli AM, Peacock TP, Thorne LG, Harvey WT, Hughes J, Covid-Genomics UK Consortium, et al. SARS-CoV-2 variant biology: immune escape, transmission and fitness. Nat Rev Microbiol 2023; 21:162-77.

22. Menegale F, Manica M, Zardini A, Guzzetta G, Marziano V, d’Andrea V, et al. Evaluation of waning of SARS-CoV-2 vaccine-induced immunity: a systematic review and meta-analysis. JAMA Netw Open 2023; 6:e2310650.

23. Reuschl AK, Thorne LG, Whelan MVX, Ragazzini R, Furnon W, Cowton VM, et al. Evolution of enhanced innate immune suppression by SARS-CoV-2 Omicron subvariants. Nat Microbiol 2024; 9:451–63.

24. Desikan R, Linderman SL, Davis C, Zarnitsyna VI, Ahmed H, Antia R. Vaccine models predict rules for updating vaccines against evolving pathogens such as SARS-CoV-2 and influenza in the context of pre-existing immunity. Front Immunol 2022; 13:985478.

