## Supplemental file for "Adjuvant Recombinant SARS-CoV-2 Spike Protein Vaccine Immunogenicity After Prior mRNA Vaccine Doses"

### SUPPLEMENTAL DATA

#### Statistical methodology

*Analysis of the primary/secondary immunogenicity objectives: nAb*

As the primary/secondary objectives of characterizing nAb response to ancestral strain Novavax vaccine administered as an additional dose after licensed mRNA vaccines and a previous ancestral strain Novavax vaccine dose, the derived/calculated endpoints of nAb response for participants who received ancestral strain vaccine included GMT, $GMFR_{Post/Pre}$, and $GMFR_{312/307}$ and SCR. nAb GMT calculated as the antilog of the mean of the log-transformed nAb titers, along with its corresponding two-sided 95% CIs, were obtained by exponentiating the corresponding log-transformed means and their 95% CIs. $GMFR_{Post/Pre}$ calculated as the within-group ratio of post-vaccination nAb GMT at Day 28 to pre-vaccination nAb GMT at baseline (Day 0), along with its 95% CIs, were estimated using paired *t*-distribution. $GMR_{312/307}$ was defined as the ratio of nAb geometric mean titers (GMT) 28 days after a dose with ancestral strain Novavax vaccine in Study 2019nCoV-312 versus 28 days after a dose with ancestral strain Novavax vaccine in Study 2019nCoV-307 among the same participants, with the corresponding 95% CIs conducted using paired t distribution. SCR was defined as the proportion of participants who achieve seroconversion equivalent to a ≥4-fold increase from baseline in nAb titers at Day 28 if the baseline value is equal to or above LLOQ or at least a 4-fold rise from LLOQ if the baseline value is lower than LLOQ. SCR in nAb GMTs with corresponding two-sided exact binomial 95% CIs was calculated using the Clopper–Pearson method. Two-sided 95% CIs of the difference in SCRs ($SCR_{312}$ – $SCR_{307}$) were based on the method of CIs for the difference in two correlated proportions by Tango (details in SAP Section 11.6). SCR differences were compared for nAb titers 28 days after a dose of ancestral strain Novavax vaccine in Study 2019nCoV-312 relative to Day 0 versus 28 days after a dose of ancestral strain Novavax vaccine in Study 2019nCoV-307 relative to Day 0 among the same participants.

Non-inferiority was demonstrated if the LB of $GMR_{312/307}$ of nAb titers was higher than 0.67 and the LB of the 95% exact CIs for the difference in SCRs in nAb titers (SCR312 – SCR307) were higher than −10%. Superiority was demonstrated if the LB of $GMR_{312/307}$ of nAb titers was higher than 1.0 after non-inferiority was successfully demonstrated.

*Conversion calculations for subsequent dose comparison among studies*

Since the pseudovirus-based nAb response against ancestral Wuhan strain at 4 different visits, including pre- and post-dose in the previous study (2019nCoV-307) and pre- and post-dose in the current study (2019nCoV-312), were tested using the similar pseudovirus-based neutralization assay by two different labs (ie, Monogram Biosciences [San Francisco, CA] and Novavax Clinical Immunogenicity [Gaithersburg, MD]), the below conversions were applied to the individual values to convert the value in units of $ID_{50}$ to the value in standard units of IU/mL.

2019nCoV-307 (Monogram): $ID_{50}$ x 0.1458 = IU/mL.

2019nCoV-312 (Novavax Clinical Immunogenicity): $ID_{50}$ x 0.214 =IU/mL.

*Analysis of the secondary immunogenicity endpoints: IgG and hACE2 receptor binding inhibition*

As the secondary objectives were demonstrating the noninferior immunogenicity of the Novavax vaccine as a dose in the current study versus a dose of the ancestral strain vaccine in the previous study following mRNA vaccines, the derived/calculated endpoints of IgG response for participants receiving the ancestral strain vaccine included IgG GMEU, $GMFR_{Post/Pre}$, $GMR_{312/307}$, SCR, and difference in SCRs, and those of hACE2 receptor-binding inhibition included hACE2 GMT, $GMFR_{Post/Pre}$, $GMR_{312/307}$, SCR, and difference in SCRs, which were calculated in a similar manner to nAb response, as stated above. The non-inferiority criteria (LB of $GMR_{312/307}$ 95% CIs being >0.67 and LB of 95% CIs for the difference in SCRs being −10%) and the superiority criterion (LB of $GMR_{312/307}$ 95% CIs being >1.0) were applied on IgG response and hACE2 receptor-binding inhibition.

As the secondary objectives were characterizing the cross-reaction of neutralizing and IgG antibodies induced by the ancestral strain vaccine (Wuhan) to a more recent SARS-CoV-2 variant (Omicron BA.5), the derived/calculated endpoints of nAb response and IgG response for participants receiving the Omicron BA.5 vaccine, including GMT/GMEU, $GMFR_{Post/Pre}$, $GMR_{312/307}$, SCR, and difference in SCRs, were calculated in a similar manner to nAb/IgG response for participants receiving the ancestral strain vaccine, as stated above.

*Analysis of the exploratory immunogenicity endpoints: JN.1 and BA.2.86*

The same statistical methods for the calculation of GMT, $GMFR_{Post/Pre}$, and SCR were applied for the analysis of nAb response against emerging variants of SARS-CoV-2 (JN.1 and BA.2.86) to evaluate immune responses developed based on the assays used as the exploratory endpoints.

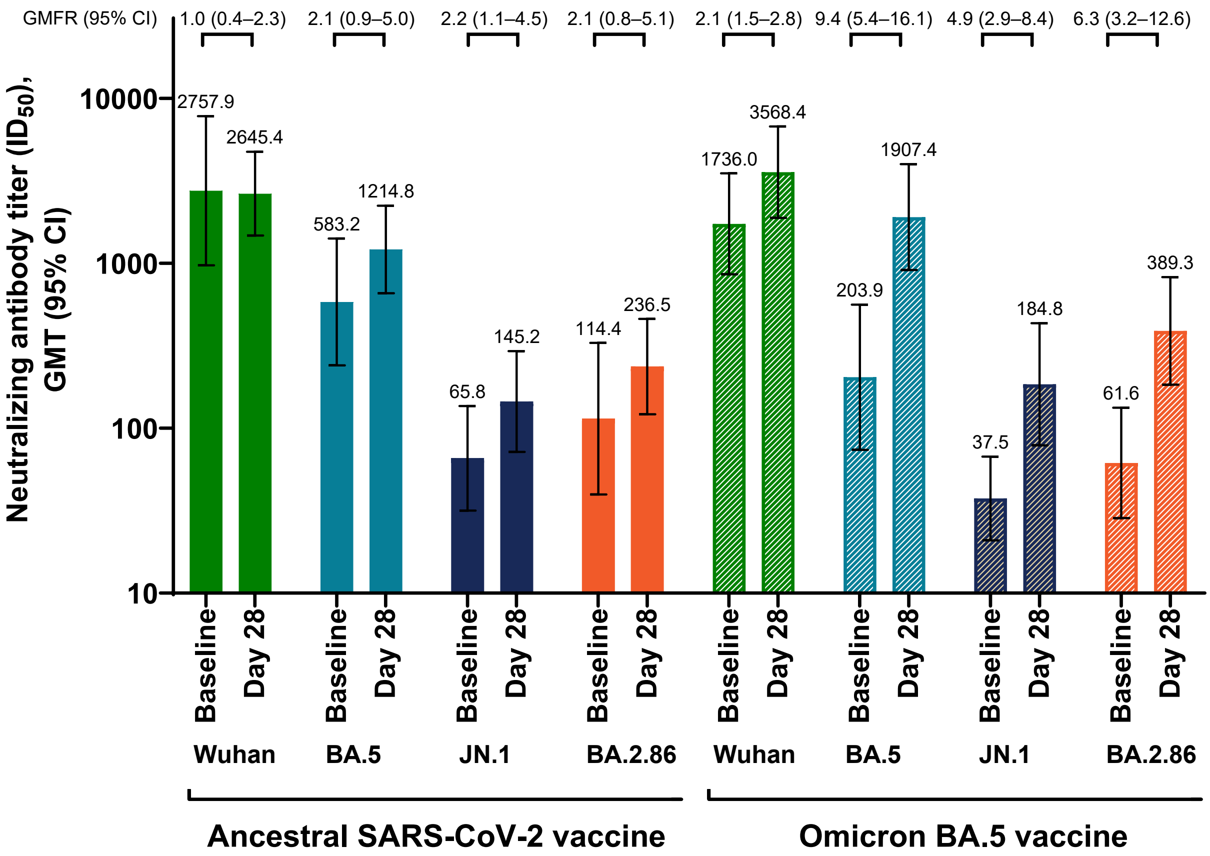


**FIG S1.** Neutralizing antibodies for various SARS-CoV-2 strains after a dose of either the ancestral SARS-CoV-2–based vaccine or the Omicron BA.5–based vaccine in the current study.

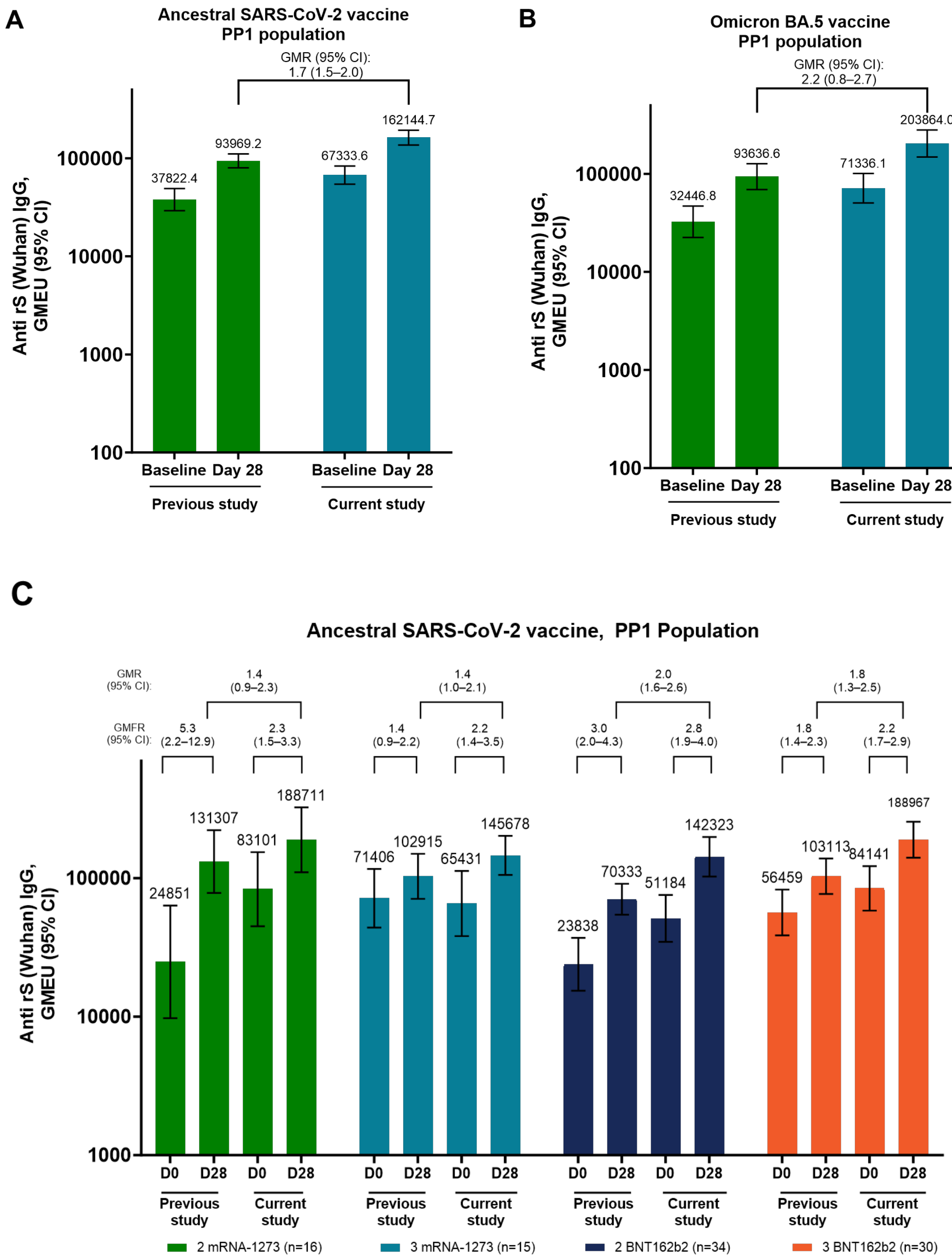
A
Ancestral SARS-CoV-2 vaccine
PP1 population
GMR (95% CI):
1.7 (1.5–2.0)
Anti rS (Wuhan) IgG,
GMEU (95% CI)
37822.4
93969.2
67333.6
162144.7
Baseline
Day 28
Previous study
Current study
B
Omicron BA.5 vaccine
PP1 population
GMR (95% CI):
2.2 (0.8–2.7)
32446.8
93636.6
71336.1
203864.0
C
Ancestral SARS-CoV-2 vaccine, PP1 Population
GMR (95% CI):
1.4 (0.9–2.3)
1.4 (1.0–2.1)
2.0 (1.6–2.6)
1.8 (1.3–2.5)
GMFR (95% CI):
5.3 (2.2–12.9)
2.3 (1.5–3.3)
1.4 (0.9–2.2)
2.2 (1.4–3.5)
3.0 (2.0–4.3)
2.8 (1.9–4.0)
1.8 (1.4–2.3)
2.2 (1.7–2.9)
24851
131307
83101
188711
71406
102915
65431
145678
23838
70333
51184
142323
56459
103113
84141
188967
D0
D28
Previous study
Current study
2 mRNA-1273 (n=16)
3 mRNA-1273 (n=15)
2 BNT162b2 (n=34)
3 BNT162b2 (n=30)

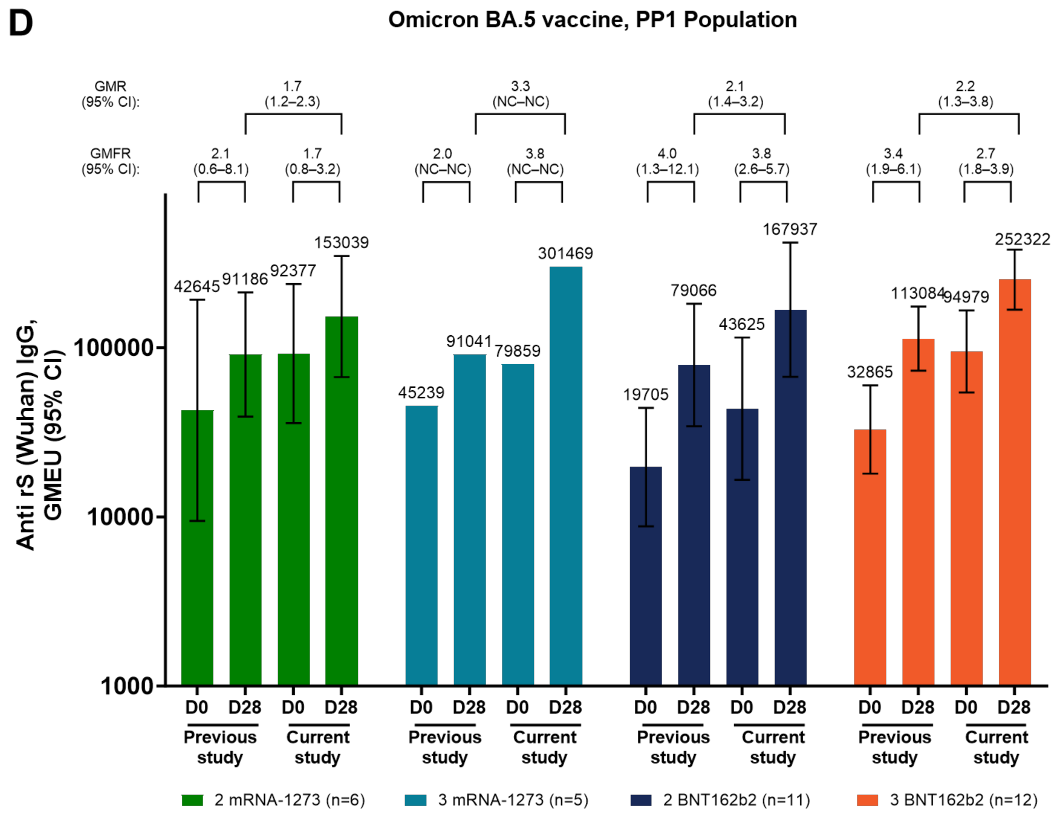


**FIG S2.** Anti-rS (Wuhan) IgG after a dose in the current study with either the ancestral SARS-CoV-2 vaccine or the Omicron BA.5 vaccine (PP1 population). Data are shown for (A) vaccination with NVX-CoV2373; (B) vaccination with NVX-CoV2540; (C) vaccination with NVX-CoV2373, assessed by prior vaccination; and (D) vaccination with NVX-CoV2540, assessed by prior vaccination. NC, not calculated because of small n. Participants who received an initial primary series of BNT162b2, 1 mRNA-1273 dose, and 1 prior NVX-CoV2373 dose before the previous study are not shown in part C because of small sample sizes (n = 2). There were no participants who received an initial primary series of mRNA-1273, 1 BNT162b2 dose, and 1 prior NVX-CoV2373 dose before the previous study.

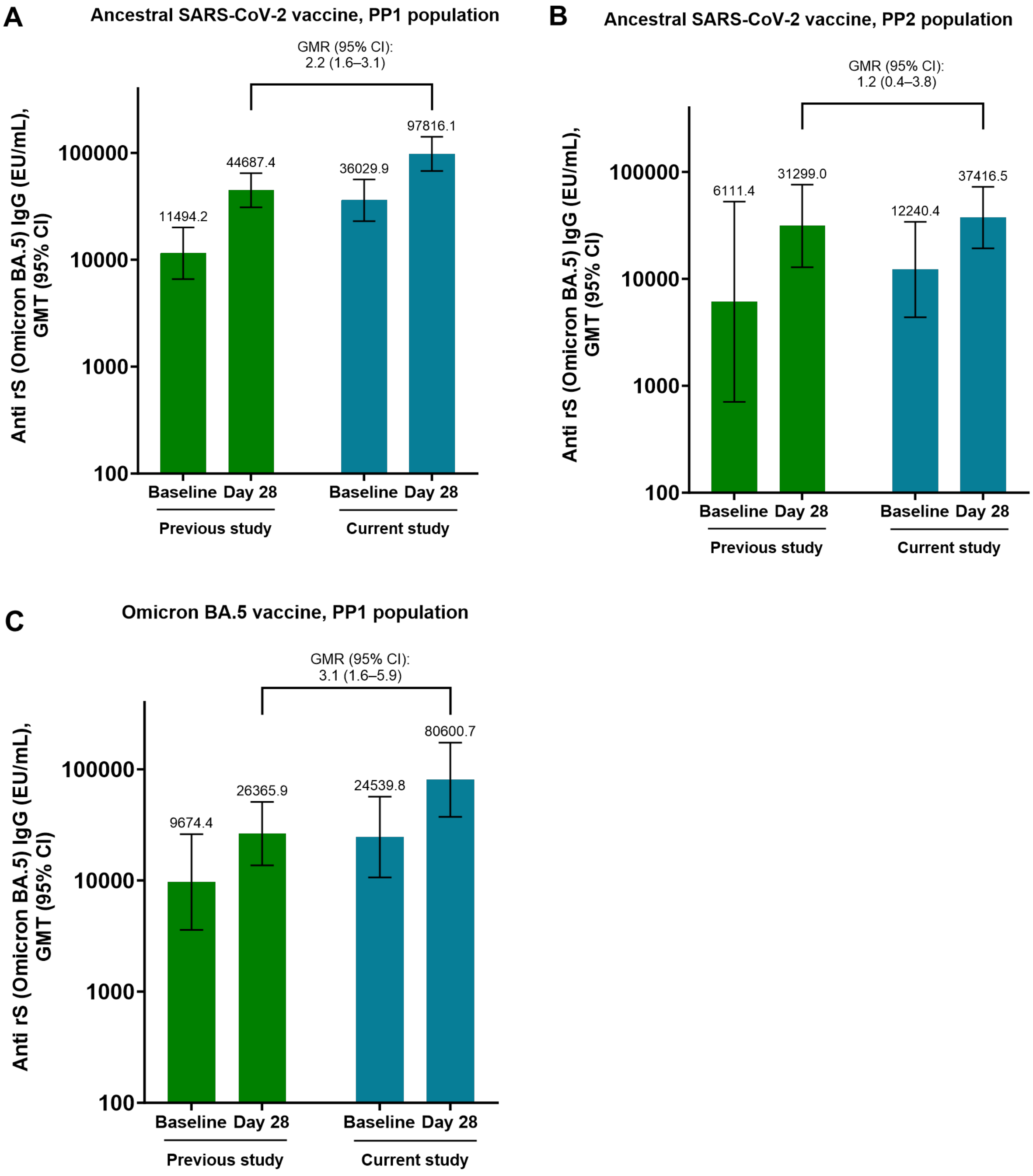


**FIG S3.** Anti-rS (Omicron BA.5) IgG after a dose in the current study with ancestral SARS-CoV-2 vaccine for the (A) PP1 population or (B) the PP2 population, or after a dose in the current study with Omicron BA.5 vaccine for the (C) PP1 population. The PP2 population for the Omicron BA.5 vaccine is not shown because of a small sample size (n = 1).

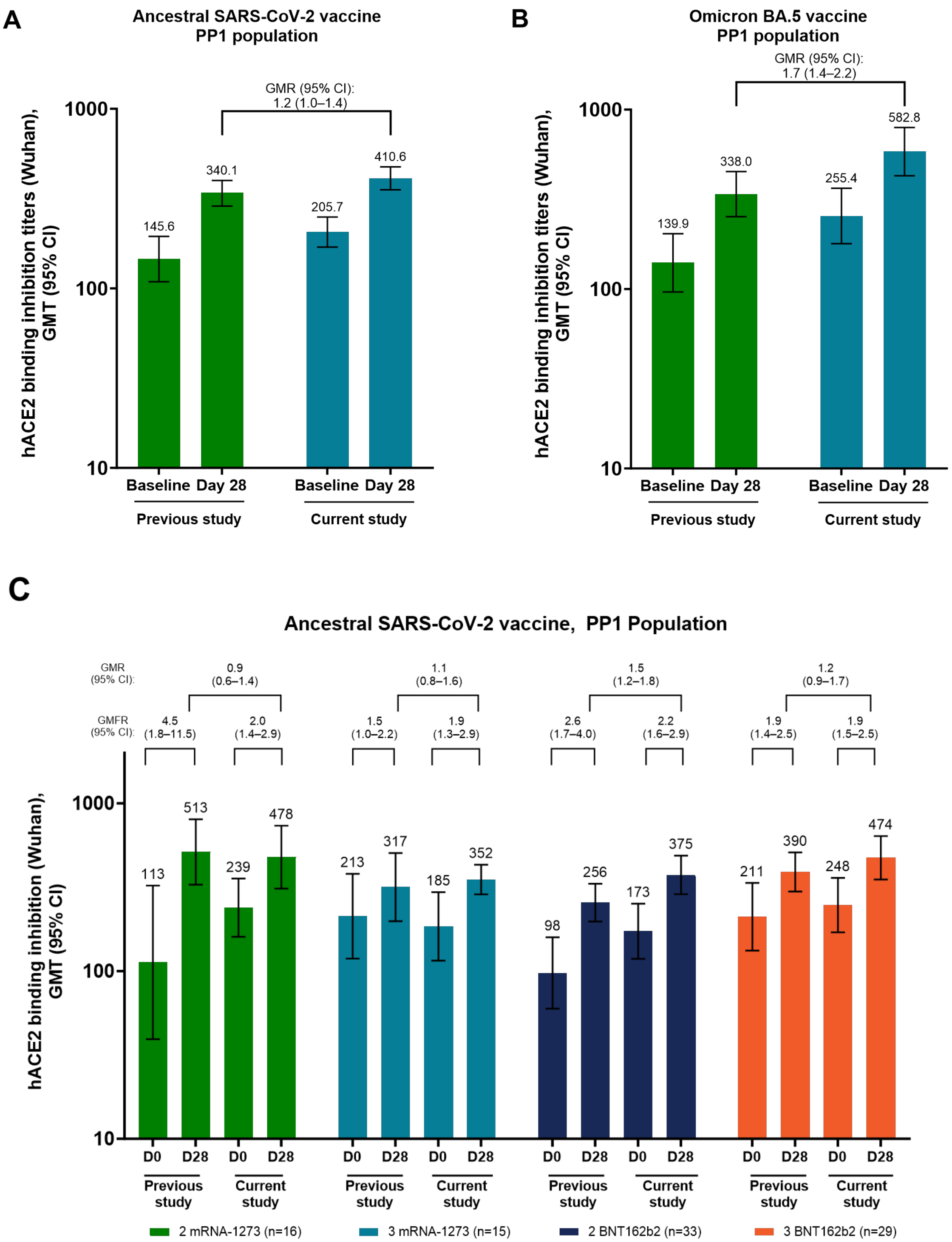

A
Ancestral SARS-CoV-2 vaccine
PP1 population
GMR (95% CI):
1.2 (1.0–1.4)
hACE2 binding inhibition titers (Wuhan),
GMT (95% CI)
1000
100
10
145.6
340.1
205.7
410.6
Baseline
Day 28
Baseline
Day 28
Previous study
Current study
B
Omicron BA.5 vaccine
PP1 population
GMR (95% CI):
1.7 (1.4–2.2)
hACE2 binding inhibition titers (Wuhan),
GMT (95% CI)
1000
100
10
139.9
338.0
255.4
582.8
Baseline
Day 28
Baseline
Day 28
Previous study
Current study
C
Ancestral SARS-CoV-2 vaccine, PP1 Population
GMR
(95% CI):
0.9
(0.6–1.4)
1.1
(0.8–1.6)
1.5
(1.2–1.8)
1.2
(0.9–1.7)
GMFR
(95% CI):
4.5
(1.8–11.5)
2.0
(1.4–2.9)
1.5
(1.0–2.2)
1.9
(1.3–2.9)
2.6
(1.7–4.0)
2.2
(1.6–2.9)
1.9
(1.4–2.5)
1.9
(1.5–2.5)
hACE2 binding inhibition titers (Wuhan),
GMT (95% CI)
1000
100
10
113
513
239
478
213
317
185
352
98
256
173
375
211
390
248
474
D0
D28
D0
D28
D0
D28
D0
D28
D0
D28
D0
D28
D0
D28
D0
D28
Previous
study
Current
study
Previous
study
Current
study
Previous
study
Current
study
Previous
study
Current
study
2 mRNA-1273 (n=16)
3 mRNA-1273 (n=15)
2 BNT162b2 (n=33)
3 BNT162b2 (n=29)

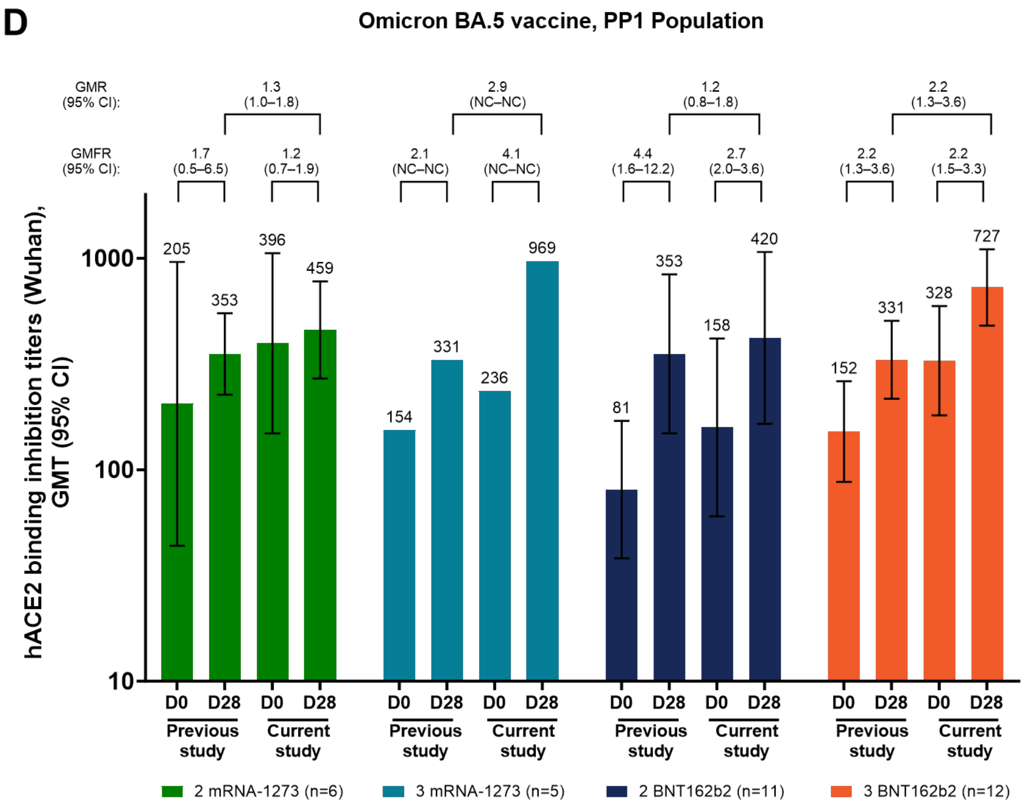


**FIG S4.** hACE2 receptor binding inhibition titers (Wuhan) after a dose in the current study with either ancestral SARS-CoV-2 vaccine or Omicron BA.5 vaccine (PP1 population). Data are shown for (A) vaccination with ancestral SARS-CoV-2 vaccine; (B) vaccination with Omicron BA.5 vaccine; (C) vaccination with ancestral SARS-CoV-2 vaccine, assessed by prior vaccination; and (D) vaccination with Omicron BA.5 vaccine, assessed by prior vaccination. NC, not calculated because of small n. For parts C and D, participants who received an initial primary series of BNT162b2, 1 mRNA-1273 dose, and 1 prior NVX-CoV2373 dose before the previous study are not shown because of small sample sizes (n = 2). There were no participants who received an initial primary series of mRNA-1273, 1 BNT162b2 dose, and 1 prior NVX-CoV2373 dose before the previous study.

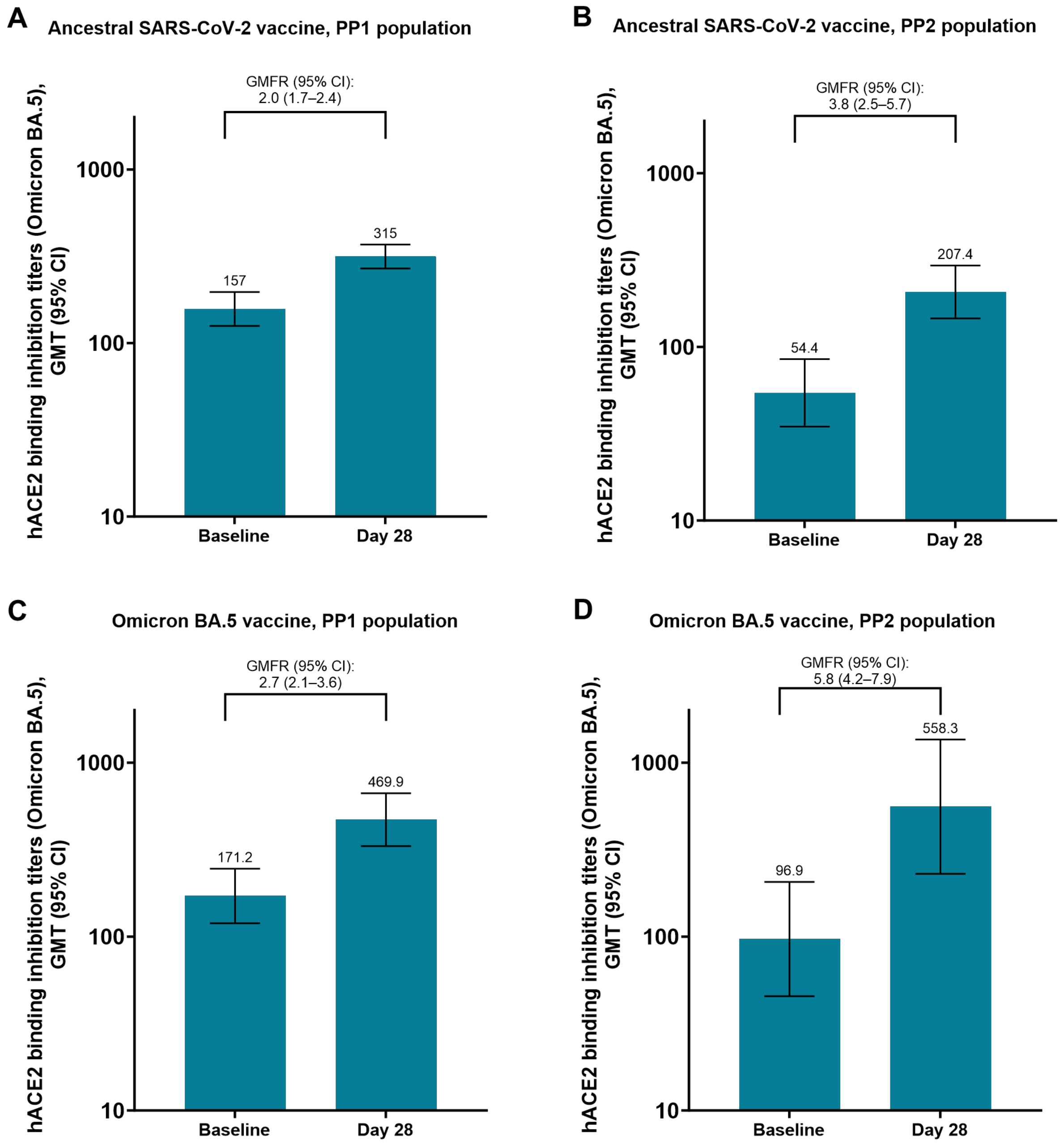


**FIG S5.** hACE2 receptor binding inhibition titers (Omicron BA.5) after a dose in the current study with ancestral SARS-CoV-2 vaccine for the (A) PP1 population or (B) PP2 population, or after a dose in the current study with Omicron BA.5 vaccine for the (C) PP1 population or (D) PP2 population.
